# School-based opportunities to improve student healthy eating, physical activity, and prevent obesity: An inventory of evidence-supported options aligned to best practice guideline recommendations

**DOI:** 10.1101/2025.06.08.25329223

**Authors:** Rebecca K Hodder, Kate M O’Brien, Nicole Nathan, Sasha Lorien, Peter Butler, Emma Ainsworth, Courtney Barnes, Ashley Blowes, Luke Wolfenden

## Abstract

**Background:** Schools are a valuable setting for the implementation of effective interventions to improve healthy eating, physical activity, sedentary behaviour and prevent overweight and obesity in children recommended by global guidelines. Robust evidence from systematic reviews, considered the gold standard source of evidence to inform public health policy and practice decisions, suggests school-based interventions produce modest improvement however effects across trials are heterogenous. To date no systematic reviews have sought to comprehensively consolidate the global evidence to identify the effectiveness of specific healthy eating and physical activity components of school-based interventions as aligned to current guidelines.

**Objective:** To 1) develop an inventory of global guideline-aligned recommendations of school-based healthy eating, physical activity, sedentary behaviour intervention componentry targeting children 5 to 12 years, and 2) explore their effectiveness via consolidation, and secondary data analysis, of existing systematic review evidence.

**Methods:** A mixed methods study was conducted, which incorporated: 1) an umbrella review of the global systematic review evidence to develop an inventory of individual school-based healthy eating, physical activity, and sedentary behaviour intervention components; and 2) consolidation of existing systematic review evidence and secondary data analysis of primary studies to explore the effectiveness of individual intervention components.

**Results:** Of 8745 records, 228 full texts were screened against eligibility criteria which identified 12 eligible systematic reviews of which eight isolated the effects of individual intervention components. Forty-nine individual healthy eating (n=26), physical activity or sedentary behaviour (n=22), or other obesity prevention (n=1) intervention components were included in the inventory. Beneficial components were identified in each of the seven recommended opportunities for multicomponent school-based interventions: healthy eating in the classroom (e.g. curriculum, food growing experiences, fruit and vegetable breaks), healthy food available at school (e.g. strategies to encourage selection of healthier foods, free cooled water, free fruit and vegetables), healthy food brought to school (e.g. information on healthy food to bring to school), healthy eating outside of school/involving families (e.g. interactive family programs), physical activity in the classroom (e.g. short activity breaks, curriculum, increased intensity/quality of physical education (PE) classes), physical activity outside of the classroom/during break times (e.g. teacher/student led organised physical activity at break times), physical activity outside of school/involving families (e.g. interactive family programs, information to parents on benefits of physical activity).

**Conclusions:** The study findings demonstrate the breadth of components that have been trialed as part of multi-component school-based interventions globally and identified a range of components that are likely beneficial in addressing obesity-related behaviours and could be prioritised in future school-based interventions.

## BACKGROUND

Poor diet, physical inactivity and obesity are responsible for a considerable burden of disease internationally,^1–4^ and the existence of these risk factors in childhood increases the risk of future chronic disease.^5^ As such, developing and implementing effective interventions targeting children is recommended to improve child health and mitigate future health burdens.^1^ One of the most valuable settings to do so is schools, as they provide centralised and universal access to students, for prolonged periods during a critical stage in their development.^6^

Robust evidence to inform policy and practice decision making is a cornerstone of evidence-based public health paradigm^7^ as it improves the impact of investments of scarce resources in public health initiatives. Rigorous systematic reviews are recommended as the ‘gold standard’ source of evidence to assess and or compare the effectiveness of different public health interventions, as they systematically search for, synthesise and appraise all relevant trial evidence.^8^ A large number of systematic reviews have been published describing the effectiveness of school-based interventions to improve healthy eating and physical activity.^9–11^ Broadly, these reviews suggest that school-based interventions produce modest improvements in these risk factors for students, however, effects across trials are heterogeneous.

Despite the number of systematic reviews available, policy makers and practitioners are critical of their utility in supporting decisions regarding the development or selection of interventions suitable for their context.^12–14^ Audits of systematic reviews in child obesity prevention, for example, have found that few (7%) have co-authors from policy-based agencies, or discuss the policy implications of their findings (29%).^15^ Others have noted concerns regarding the likely generalisability of international literature across contexts.^14,16^ This is particularly salient for school-based programs promoting healthy eating, physical activity, or obesity prevention behaviours, as school systems vary considerably across jurisdictions. Such differences are evident even between high-income countries. For example, in the United States and Canada a primary source of food consumed in schools comes from school cafeterias, while in Australia and New Zealand, food consumed at schools is primarily brought from home and packed in a lunchbox.^17,18^ Such differences necessitate different approaches to improving healthy eating behaviours in these settings.

Several frameworks and guidelines have been published to support effective promotion of student healthy eating, physical activity, and obesity prevention in this setting. These recommend comprehensive, multi-component approaches. For example, the World Health Organisation Health Promoting Schools framework suggests that approaches to improve student health that are holistic and include strategies targeting the school curriculum, ethos and/or environment and that engage families and communities can be effective in improving weight related behaviours.^19^ Other best practice guidelines in Australia and internationally recommend the implementation of intervention components that target opportunities to improve healthy eating and physical activity across the school day, including within the classroom, during breaks, and at home.^20–22^

To our knowledge, however, no systematic review has comprehensively consolidated global evidence on the effectiveness of specific healthy eating and physical activity components of school-based interventions for children 5 to 12 years, as tested to date and aligned to current guidelines.^23–27^ Evidence regarding the benefit of individual healthy eating and physical activity intervention components is critical to guide the development of more efficient and effective school-based initiatives. For example, a range of different strategies could be employed as part of a multicomponent intervention to improve the availability or selection of healthy foods at school, such as changes to food provided or sold on site, the introduction of fruit and vegetable breaks for students, or introducing labelling nudges that support healthy food selection.^9–11^ An inventory of the types of strategies that have been employed, and an assessment of their effectiveness would aid the selection of more potent and contextually relevant intervention components. The absence of such information and analyses is an important gap in the literature.

In this context, the aim of this study is to 1) develop an inventory of global guideline-aligned recommendations of school-based healthy eating, physical activity intervention componentry targeting children 5 to 12 years, and 2) explore their effectiveness via consolidation, and secondary data analysis, of existing systematic review evidence.

## METHODS

A mixed methods study was conducted, which incorporated: 1) an umbrella review of the global systematic review evidence to develop an inventory of individual school-based healthy eating and physical activity intervention components; and 2) consolidation of systematic review evidence and secondary data analysis of primary studies to explore the effectiveness of individual intervention components.

### 1. Umbrella review of the global systematic review evidence to develop an inventory of individual school-based healthy eating and physical activity intervention components

We conducted an umbrella review of the global review evidence to develop an inventory of individual healthy eating and physical activity components that have previously been trialed. The review was conducted in line with Cochrane recommendations for umbrella reviews.^28^

#### Study inclusion criteria

We sought systematic reviews published in the last five years that implemented a comprehensive search strategy and synthesised the effects of randomised controlled trials (RCTs) (individual or cluster) on healthy eating, physical activity or obesity prevention interventions targeting healthy eating and/or physical activity in school-based interventions for children aged 5 to 12 years. Eligible systematic reviews had to report a synthesis of RCTs, with or without meta-analysis, and could be either a review of primary studies or an umbrella review of systematic reviews of RCTs. Based on Cochrane guidelines, we defined systematic and umbrella reviews high-quality if they included explicit reproducible methodology i.e., included a comprehensive search with explicit criteria for including or excluding studies, and acceptable methods for assessing the quality of included studies (i.e. using an existing tool or reporting the criteria used to make assessments on the quality of the included studies). Reviews that did not specifically synthesise the effects of school-based RCTs (either in meta-analysis or narratively) were excluded. Reviews including a combination of RCTs and other study designs (e.g. non-randomised trials) were included if the results were reported separately for RCTs. Eligible review outcomes for each outcome category included any measure of healthy eating, physical activity and child weight status

#### Search strategy and review selection

A Medline search was conducted from inception up to end 2023 to identify potentially eligible systematic or umbrella reviews of systematic reviews published in the last 5 years that synthesised the effects of school-based healthy eating, physical activity or obesity prevention interventions on healthy eating, physical activity, or child weight (Appendix A).

#### Review selection

Records were de-duplicated, uploaded and screened using Covidence software.^29^ One reviewer conducted an initial screening of titles only to discard irrelevant titles. Pairs of review authors then independently screened titles, abstracts, and full text articles of all identified reviews in duplicate for eligibility. Disagreements for title, abstract, and full text screening were resolved by discussion and consensus, or by consultation with a third review author. To develop the inventory, we extracted details of primary studies included in the most recent, high-quality and comprehensive reviews targeting each of the following outcome categories: child healthy eating; child physical activity; and child obesity prevention. We defined a comprehensive and high-quality review as one that adopted methods consistent with the Cochrane handbook. We prioritised systematic reviews of primary studies (RCTs) that synthesised findings using meta-analysis, and if no such reviews were available, we included umbrella reviews of systematic reviews that included RCTs.

#### Data extraction for inventory development

Interventions and their components of each primary study extracted from the selected reviews were independently coded by pairs of health promotion practitioners with experience in the implementation of healthy eating and physical activity interventions in schools. An inventory of each discrete healthy eating or physical activity intervention component was iteratively developed during this process, which included consensus-based definitions. Discrepancies between coders were resolved via discussion or via consultation with a third reviewer.

Intervention components were then coded against current recommendations of existing International^19–21,30–43^ and Australian^22,44–46^ best-practice guidelines regarding school-based healthy eating, physical activity and obesity prevention strategies. To do so, we reviewed and consolidated current recommendations into a framework of seven commonly recommended school-based opportunities to improve healthy eating or physical activity, and one other opportunity targeting child weight status. The framework is described in table 1 and includes citation details demonstrating concordance of the framework with current guidelines (Appendix B).

**Table 1.** Consolidated framework of school-based healthy eating and physical activity opportunities

| Opportunities for healthy eating and physical activity |  | Examples |
| --- | --- | --- |
| Healthy eating | 1. Healthy eating in the classroom<sup>20-22,30-32,34-36,38,39,41-43,46</sup> | Healthy eating lessons, breaks for fruit and vegetable consumption during class |
|  | 2. Availability of healthy food at school<sup>20-22,30-32,34-36,38,41-44,46</sup> | Healthy canteen strategies, incentives for healthy food choices |
|  | 3. Healthy food brought to school<sup>35,38,42,43</sup> | Healthy lunchbox strategies |
|  | 4. Healthy eating outside of school or involving families<sup>19,21,31,34,36,38,39,41</sup> | Interactive cooking programs for students and families |
| Physical activity | 5. Physical activity in the classroom<sup>20-22,30,33-37,40-42,45</sup> | Physical activity lessons, short activity breaks during class |
|  | 6. Physical activity outside the classroom and during break times<sup>20-22,30,33,34,36,37,40,41,45</sup> | Physical activity programs, access to physical activity equipment |
|  | 7. Physical activity outside of school or involving families<sup>20,21,33,34,36,37,41</sup> | Active travel promotion, interactive physical activity programs for students and families |
| Other | 8. Body mass index assessment | Height and weight screening with or without individual reports to families |

### 2. Systematic review evidence and secondary analysis of primary studies to explore the effectiveness of individual intervention components

To assess the effectiveness of discrete intervention components, we first sought to examine the findings of systematic reviews of RCTs that reported the effects of individual school-based interventions. This involved screening the citations of the umbrella review Medline search (described above) using the same eligibility criteria to identify eligible reviews that reported the effectiveness of individual healthy eating or physical activity intervention components.

Second, to explore the effectiveness of individual intervention components that were not isolated in any eligible systematic reviews, we conducted a secondary analysis of the primary studies included in the reviews used to develop the intervention component inventory.

#### Data extraction, synthesis and analysis Systematic review evidence

Where an eligible systematic review reported the effects of intervention components in isolation (narratively or via meta-analysis), we reported this effect. We extracted and reported the full citation details and review characteristics of all reviews from which we draw findings regarding intervention component effects. Quality assessments of the included systematic reviews were assessed by two independent review authors using Assessing the Methodology Quality of Systematic Reviews tool 2 (AMSTAR2).^47^ Of the 16 tool domains, we identified the following six critical domains according to the tool guidance that weighted more heavily when rating the overall confidence in results for each review: protocol registration; adequacy of literature search; performing risk of bias assessment; appropriateness of meta-analytical method; consideration of risk of bias when interpreting the results; and assessment of publication bias. Review authors assessed each according to the tool as: ‘yes’, ‘partial yes’, ‘no’, or ‘no meta-analysis conducted’ and disagreements were resolved by discussion and consensus.

#### Secondary analysis of primary studies

The individual study characteristics and study results (healthy eating, physical activity, sedentary behaviour or weight) for each primary study in the reviews used to develop the intervention component inventory were extracted. These included estimates reported for each individual study in meta-analysis results from online repositories (where available e.g. Cochrane reviews), data from study authors, or data extracted from individual studies.

To estimate the effectiveness of multicomponent interventions that have adopted each individual component, meta-analyses were conducted where possible, using primary study data using the generic inverse variance method in a random effects model with Review Manager 5 software. We calculated mean differences or standardised mean differences, and 95% confidence intervals when two or more studies reported the same outcome. For meta-analysis, an intervention including a specific component was classified as having a beneficial impact when confidence intervals of the effect estimate did not include zero effect (i.e. did not cross 0).

Meta-analysis was not performed to assess the effects of healthy eating focused intervention components, given the substantial heterogeneity of healthy eating measures included in primary studies. Instead, we calculated the proportion of primary studies reporting statistically significant beneficial effects on one or more of the eligible outcomes that included the individual component. To aid communication of the results, a cut point of ≥75% of trials was used to indicate an overall beneficial effect.

## RESULTS

### Search results

A total of 8745 records were identified in the database search, of which 228 full texts were screened against eligibility criteria and 12 systematic reviews were included (Figure 1). This included three reviews for the intervention component inventory development (Table 2) and eight reviews that reported isolated effects of individual intervention components (Table 3).

**Figure 1.**
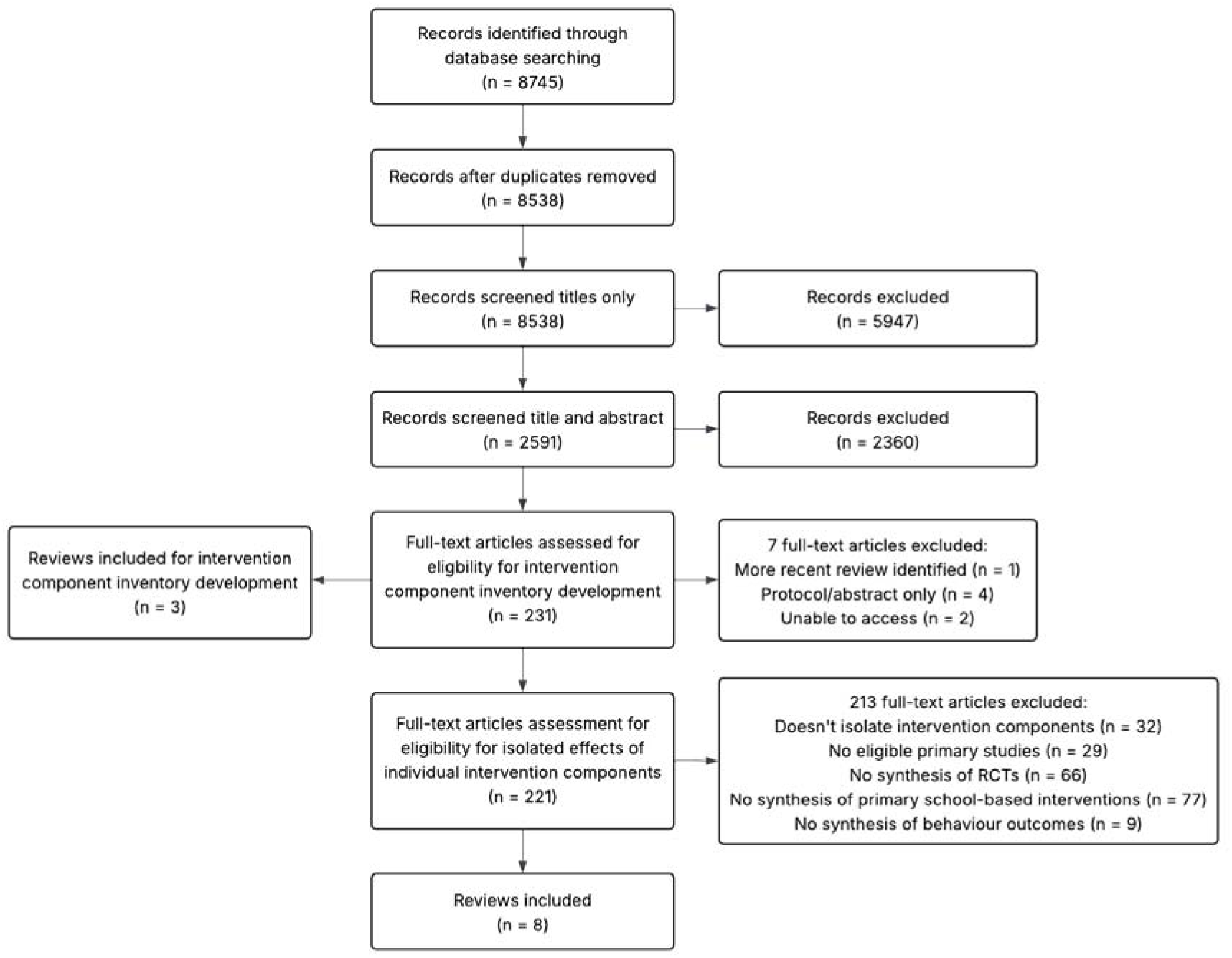
Flow diagram

**Table 2.** Characteristics of three reviews for the intervention component inventory development

| Author (year) | Eligibility criteria<br>Search date | No. of trials included | Results | Quality Assessment |
| --- | --- | --- | --- | --- |
|  |  | Years of publication |  |  |
| Hodder (2022)<sup>10</sup> | **Study design:** RCTs, CRCTs<br>**Country:** any<br>**Participants:** children aged 6 to 18 years<br>**Intervention:** child obesity prevention interventions with no restriction on intervention type, setting, or delivery personnel<br>**Comparator:** non-intervention or usual care; alternate intervention<br>**Outcome of interest:** BMI/BMI z score<br>**Search date:** June 2021 | 195 trials<br>Delivered in primary school (6-12 years): 98 trials<br>1993 to 2021 | **BMI:** SMD -0.03, 95%CI: -0.05 to -0.01; 72 trials; 114,451 participants; $I^2 = 21\%$ | High |
| Neil-Sztramko (2021)<sup>48</sup> | **Study design:** RCTs, CRCTs<br>**Country:** any<br>**Participants:** children aged 6 to 18 years<br>**Intervention:** any school-based PA program aimed to increase PA and/or fitness, with a duration of $\geq 12$ weeks<br>**Comparator:** no intervention, usual care, or a concomitant intervention<br>**Outcome of interest:** MVPA, SB<br>**Search date:** June 2020 | 89 trials<br>Delivered in primary school (6-12 years): 66 trials<br>1988 to 2020 | **MVPA:** MD 1.01 minutes/day, 95%CI: 0.08 to 1.93; 22 trials; 10,715 participants; $I^2 = 69\%$<br>**SB:** MD -3.35 minutes/day, 95%CI -9.30 to 2.60; 11 trials; 5766 participants; $I^2 = 37\%$ | High |
| O'Brien (2021)<sup>11</sup> | **Study design:** systematic reviews of RCTs, CRCTs<br>**Country:** any<br>**Participants:** children aged 6 to 18 years<br>**Intervention:** school-based nutrition interventions which aimed to improve children's dietary intake (either as primary or secondary aim of included reviewers)<br>**Comparator:** any type of comparator<br>**Outcome of interest:** dietary intake<br>**Search date:** June 2021 | Number of reviews included: 13 systematic reviews<br>2011 to 2021 | Narrative synthesis of 82 RCTs of school-based healthy eating interventions targeting children aged 6 to 12 years found some healthy eating interventions (such as nutrition education and food environment interventions) to have a positive effect of some dietary outcomes including fruit, fruit and vegetables combined, and fat intake | Quality of the 13 reviews: 2 high, 1 moderate, 3 low, 7 critically low quality |

**Abbreviations** BMI: body mass index; CRCT: cluster randomised controlled trial; CI: confidence interval; MVPA: moderate-to-vigorous physical activity; PA: physical activity; RCT: randomised controlled trial; SB: sedentary behaviour

**Table 3.** Characteristics of eight reviews that reported isolated effects of individual intervention components

| Author (year) | Eligibility criteria | No. of eligible trials included* (total trials)<br>Years of publication | Results | Quality Assessment |
| --- | --- | --- | --- | --- |
| Lonsdale (2013)<sup>49</sup> | **Study design:** RCTs, CRCTs, quasi-experimental<br>**Country:** any<br>**Participants:** children in primary or secondary school<br>**Intervention:** interventions to implement a change to usual teaching practice to increase the proportion of PE lesson time spent in MVPA<br>**Comparator:** no-intervention control<br>**Outcome of interest:** proportion of PE lesson time spent in MVPA | 14 trials (16 trials)<br>1991 to 2008 | **Proportion of PE lesson time spent in MVPA:** ES 10.37, 95%CI: 6.33 to 14.1; 13 trials; 3360 participants; $I^2 = 89\%$ (NB. 1/13 trials was quasi-experimental design) | Critically low |
| Masini (2020)<sup>50</sup> | **Study designs:** RCTs, cohort<br>**Country:** any<br>**Participants:** children aged 6 to 13 years<br>**Intervention:** active break intervention<br>**Comparator:** active learning, theoretical lesson about PA or no intervention<br>**Outcome of interest:** PA levels | 6 trials (22 trials)<br>2002 to 2019 | **MVPA:** MD 4.29, 95%CI: -0.15 to 8.74; 2 trials; 419 participants; $I^2 = 38\%$ | Critically low |
| Norris (2020)<sup>51</sup> | **Study design:** RCTs, quasi-experimental, non-randomised controlled trials<br>**Country:** any<br>**Participants:** children aged 3 to 14 years<br>**Intervention:** physically active lessons<br>**Comparator:** control<br>**Outcome of interest:** lesson-time PA, overall PA | 21 trials (42 trials)<br>2008 to 2019 | **Lesson-time PA:** SMD 2.46, 95%CI: 1.46 to 3.46; 14 trials; 4511 participants<br>**Overall PA:** SMD 0.32, 95%CI: 0.18 to 0.46; 8 trials; 4467 participants | Critically low |
| Nury (2022)<sup>52</sup> | **Study design:** CRCTs<br>**Country:** any<br>**Participants:** children aged 4 to 18 years<br>**Intervention:** whole school environment interventions (nutrition education and literacy, food preparation in school, school garden, nutrition friendly school initiatives)<br>**Comparator:** control<br>**Outcome of interest:** daily intake of fruit and vegetables, weight, BMIz score | 40 trials (51 trials)<br>2000 to 2021 | **Daily intake of fruit and vegetables:** Nutrition education/literacy: MD 33.63 g/day, 95%CI: 0.73 to 66.52; 4 trials. Nutrition friendly school initiatives (healthy canteens): MD -7.26, 95%CI: -54.96 to 40.43; 2 trials<br>**Weight (narrative synthesis):** No benefits of nutrition education/literacy and nutrition friendly school initiatives in a school setting over a control group<br>**BMIz score:** Nutrition education/literacy: MD -0.23, 95%CI: -0.34 to -0.13; 2 trials | High |
| Ohly | **Study designs:** RCTs, non-randomised controlled trials, CBA | 3 trials (40 trials) | **Fruit and vegetable intakes (narrative synthesis):** All 3 RCTs found no | Critically low |
| (2016)<sup>53</sup> | **Country:** OECD countries<br>**Participants:** children (all ages up to 18 years), school staff, family, community members (all ages)<br>**Intervention:** school gardening activities<br>**Comparator:** control groups or groups participating in alternative activities (such as nutrition education without gardening activities)<br>**Outcome of interest:** nutrient intake | 2001 to 2014 | significant changes in fruit and vegetable intake<br>**Nutrient intake (narrative synthesis):** All 3 RCTs reported statistically significant changes in nutrient intake |  |
| Peiris (2022)<sup>54</sup> | **Study design:** RCT, CRCT<br>**Country:** any<br>**Participants:** children aged 6 to 13 years<br>**Intervention:** Activity breaks interventions carried out inside the classroom<br>**Comparator:** control<br>**Outcome of interest:** MVPA, SB | 5 trials (10 trials)<br>2013 to 2021 | **MVPA:** SMD 3.55, 95%CI: 3.29 to 3.81; 3 trials; 605 participants; $I^2 = 97\%$<br>**SB:** SMD 1.01, 95%CI: -1.19 to 3.39; 2 trials; 498 participants; $I^2 = 99\%$ | Critically low |
| Pulido Sanchez (2021)<sup>55</sup> | **Study design:** RCT, CRCT<br>**Country:** any<br>**Participants:** children aged 6 to 18 years<br>**Intervention:** interventions aimed to increase PA during school recess<br>**Comparator:** control<br>**Outcome of interest:** MVPA | 6 trials (43 trials)<br>2011 to 2019 | **MVPA (narrative synthesis):** Five of six RCTs reported a significant increase; 160,846 participants | Critically low |
| Vetter (2020)<sup>56</sup> | **Study design:** RCT, CT, quasi-experimental<br>**Country:** any<br>**Participants:** primary school children<br>**Intervention:** active learning combining PA and math<br>**Comparator:** control<br>**Outcome of interest:** PA | 6 trials (11 trials)<br>2009 to 2018 | **PA (narrative synthesis):** "Of studies measuring PA with accelerometers, 4 of 5 reported significantly greater PA in the intervention group during the intervention ($p < .05$) or across the school day ($p < .01$)", 1386 participants | Critically low |

\*Eligible trials were randomised controlled trials synthesised with or without meta-analysis on healthy eating, physical activity or obesity prevention in school-based interventions for children aged 5 to 12 years.
**Abbreviations** BMI: body mass index; CBA: controlled-before-after; CT: controlled trial; CRCT: cluster randomised controlled trial; CI: confidence interval; ES: effect size; ITS: interrupted time series; MD: mean difference; MVPA: moderate-to-vigorous physical activity; OECD: organisation for economic co-operation and development; PA: physical activity; PE: physical education; RCT: randomised controlled trial; SB: sedentary behaviour; SMD: standard mean difference

### Umbrella review and inventory of individual healthy eating and physical activity intervention components

Three reviews were used to develop the intervention component inventory, one for each outcome category (Table 2). This included a review of childhood obesity prevention interventions that reported an overall effect of 71 school-based RCTs on child weight (AMSTAR2: high quality); a review of school-based physical activity interventions that reported an overall effect of 18 RCTs on moderate-to-vigorous physical activity (MVPA) and sedentary behavior (AMSTAR2: high quality); and an umbrella review of school-based healthy eating interventions that reported an overall effect of 97 RCTs from 13 systematic reviews on healthy eating outcomes (AMSTAR2: two high, one moderate, three low, and seven critically low quality). There were 155 discrete RCTs included across the three reviews, which were primarily conducted in high-income countries (83%) and published between 1996 and 2021. Further details of the critical appraisal can be found in supplementary Appendix C.

Across all interventions tested in the 155 primary studies within systematic reviews, 49 individual healthy eating (n=26), physical activity or sedentary behaviour (n=22) or other obesity prevention (n=1) intervention components were included in the inventory. Each individual intervention component was coded to one of the seven recommended opportunities for school-based interventions to improve student healthy eating or physical activity or other (Table 1). The majority of interventions (95%) tested within the primary studies were multicomponent and tested various different combinations of healthy eating and/or physical activity intervention components (range: 1-17 individual intervention components).

### Effectiveness of individual intervention components

#### Systematic review evidence

The eight included reviews reported the isolated effects of eight individual healthy eating or physical activity intervention components (Table 4) and included studies published between 1991 and 2021. Of the eight reviews, the quality according to AMSTAR2 of one review was rated as high quality, and the remainder were rates as critically low quality. Further details of the critical appraisal can be found in Appendix C.

**Table 4.**
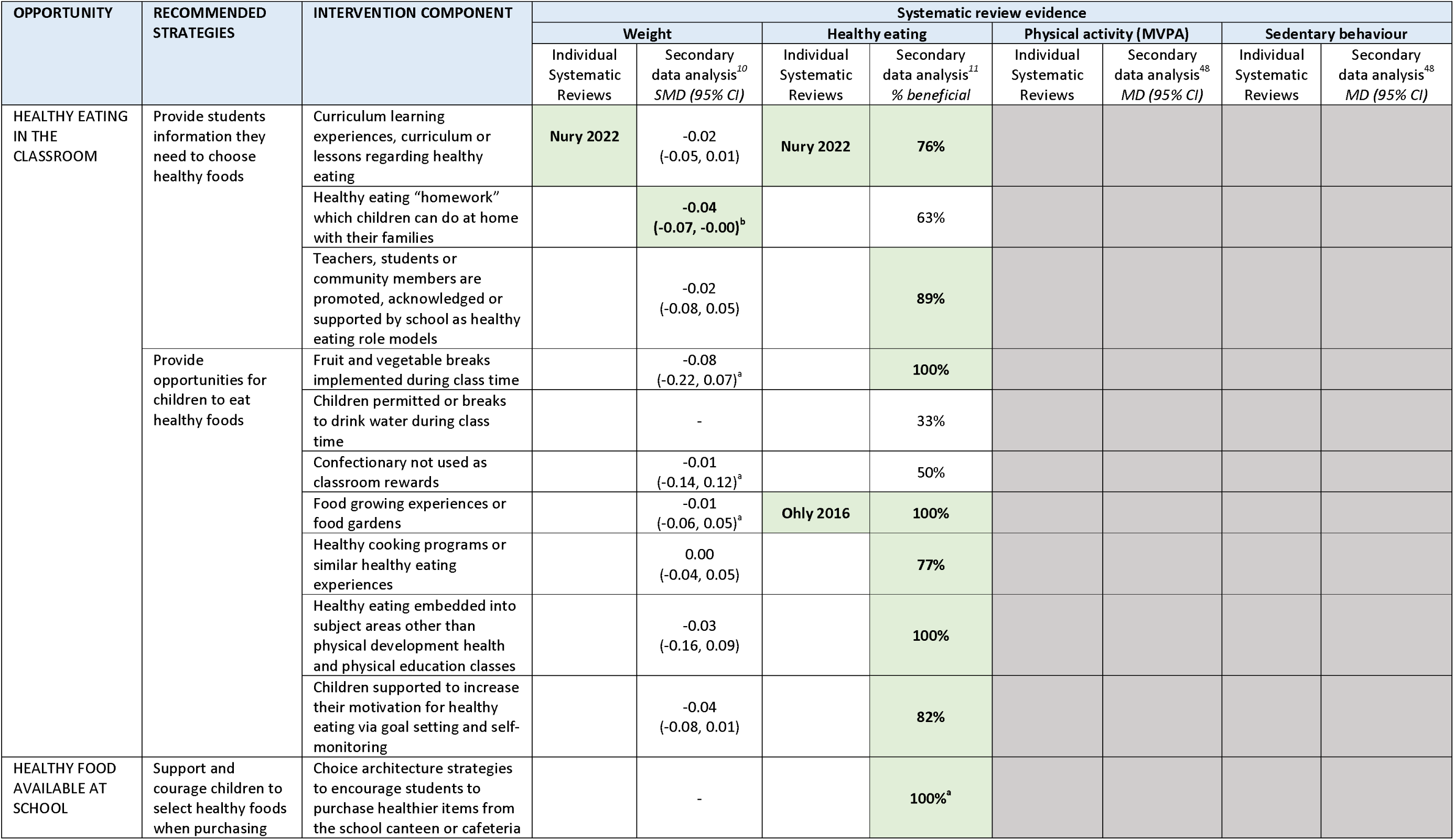
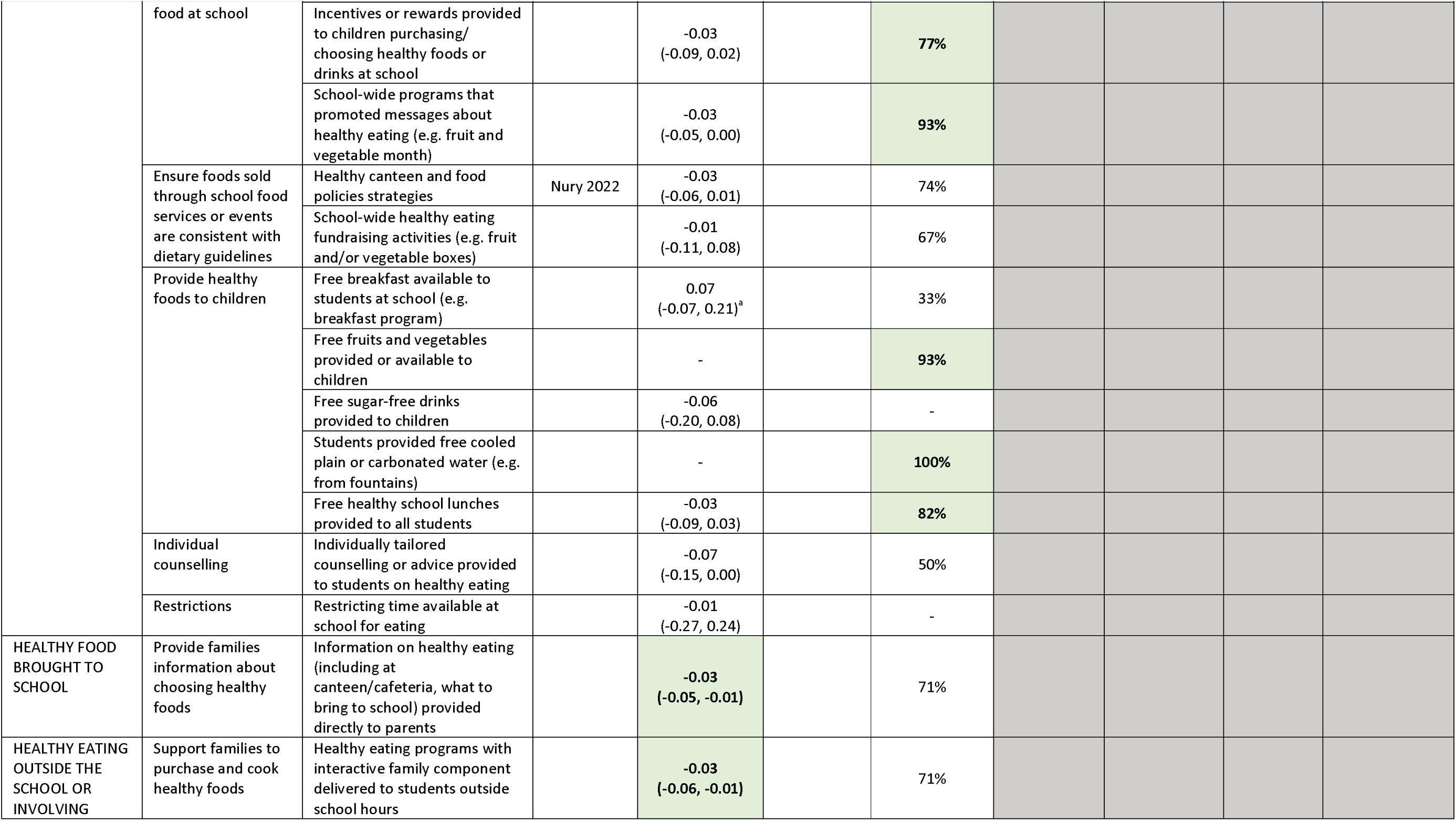
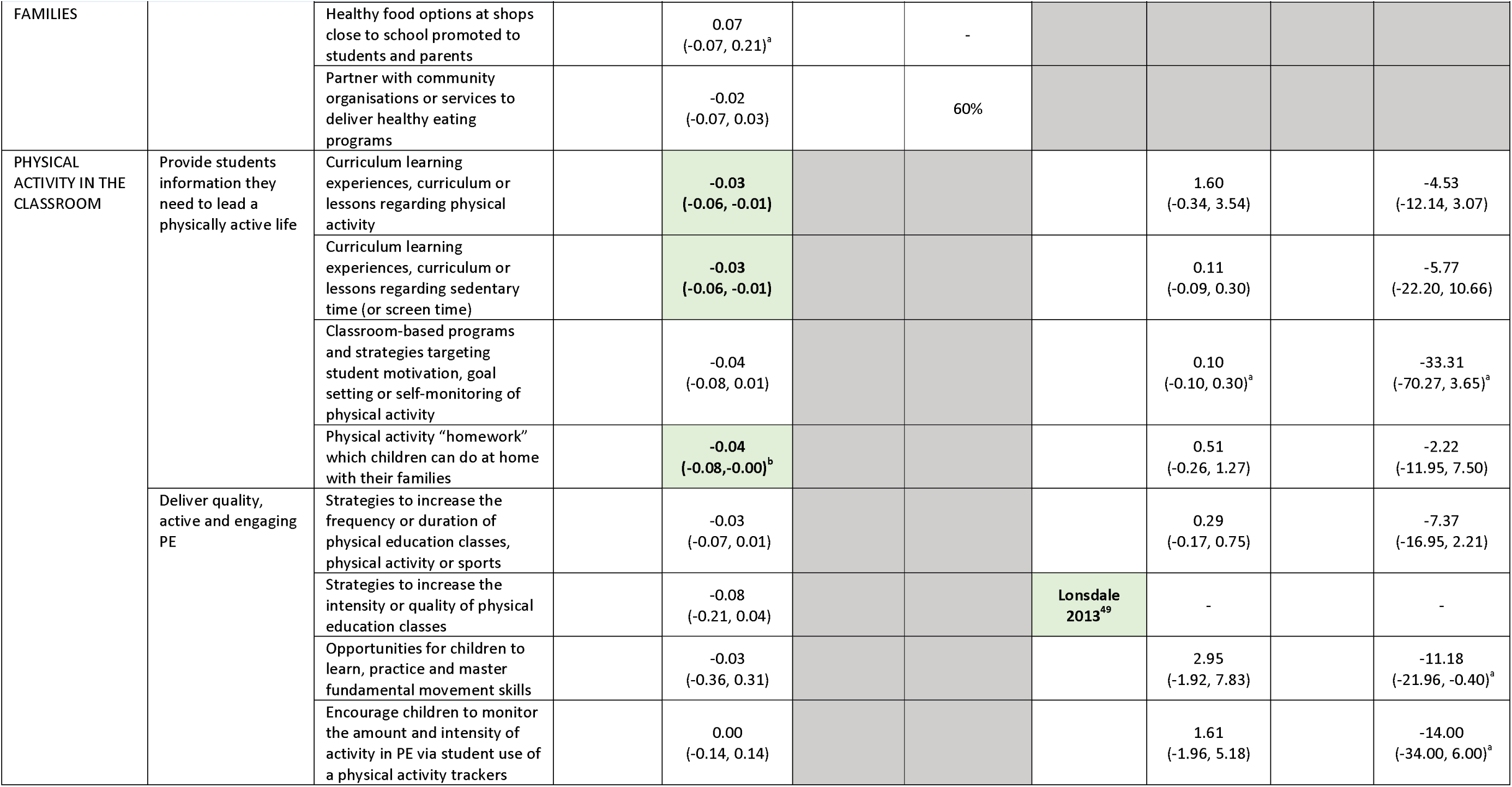
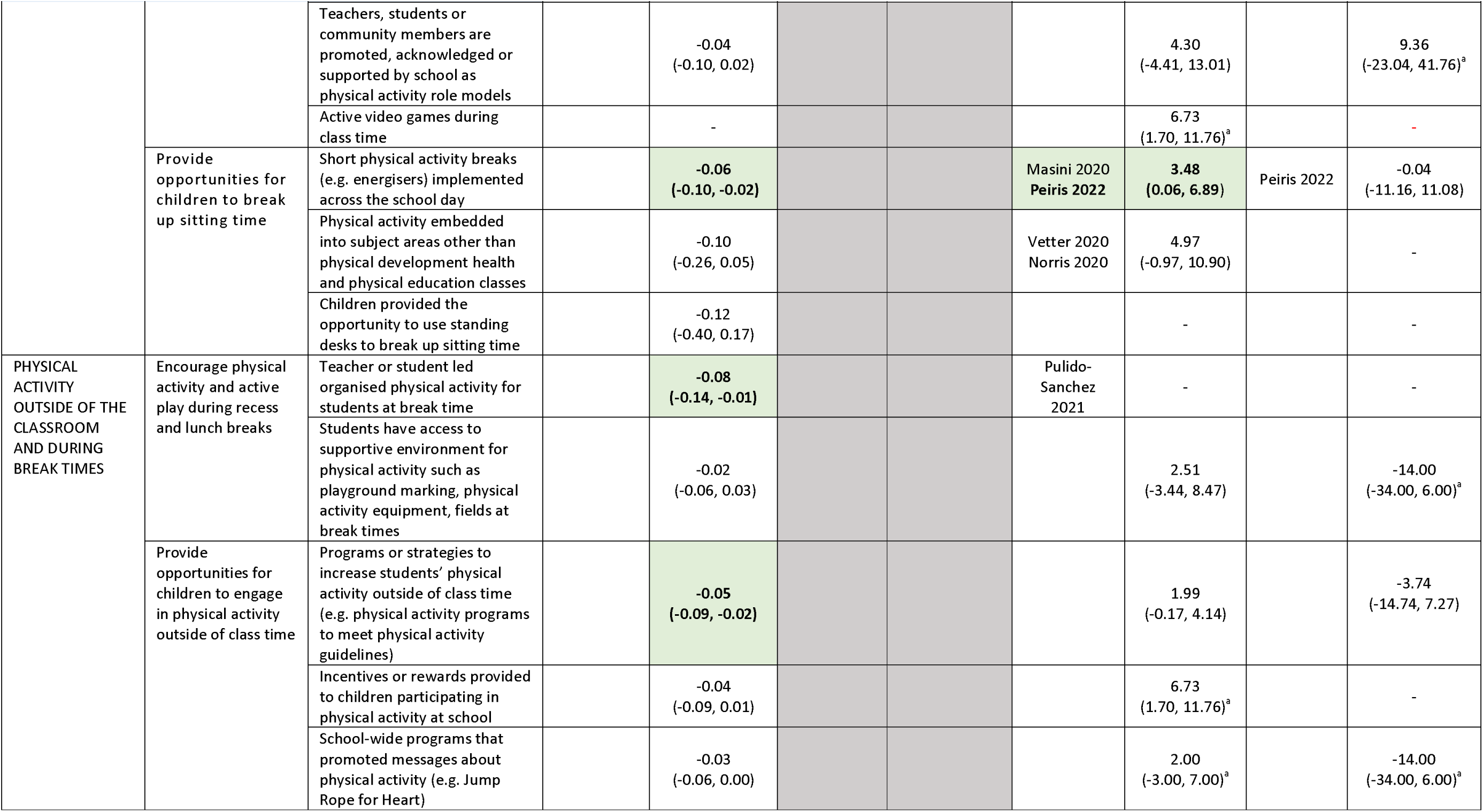
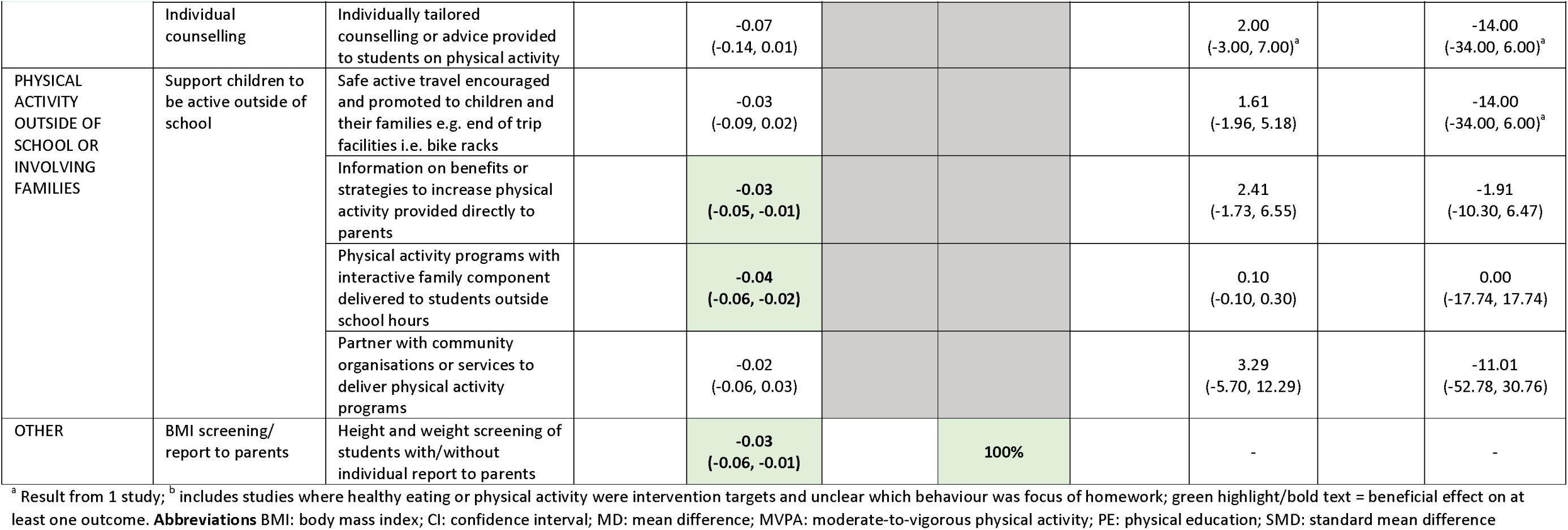
Systematic review evidence for individual healthy eating and physical activity intervention components

A beneficial impact on either healthy eating, physical activity and/or weight was found for three of the nine individual intervention components. The effects of four healthy eating components were isolated in two systematic reviews which reported a beneficial impact of “curriculum learning experiences, curriculum or lessons regarding healthy eating” on both weight and healthy eating, and “food growing experiences or food gardens” on healthy eating, but no beneficial impact of “healthy cooking programs/healthy eating experiences” or “healthy canteen policies” on weight (Table 4). The effects of five physical activity components were isolated in the other seven systematic reviews, which reported a beneficial impact of “strategies to increase the intensity or quality of physical education classes “ and “Short physical activity breaks” on MVPA, but no beneficial impact of “physical activity embedded into subject areas other than PE classes”, “teacher or student led organised physical activity at break time”, “safe active travel encouraged and promoted to children and their families” on MVPA or sedentary behaviour.

#### Secondary analysis of primary studies

Meta-analyses data of primary studies examining the effect interventions of physical activity and weight was available that contributed to one of 82 separate meta-analyses for each individual intervention component tested within the primary studies. This included separate meta-analyses examined the impact of 45 healthy eating or physical activity intervention components on weight, and 20 physical activity intervention components on MVPA, and 17 physical activity intervention components on sedentary behaviour. The proportion of studies reporting a beneficial impact on child healthy eating was calculated for 24 individual healthy eating components (no meta-analysis data of primary studies was available).

##### Healthy eating intervention components

A beneficial impact on a measure of student healthy eating or weight was reported for 16 of the 26 healthy eating intervention components.

Beneficial *‘healthy eating in the classroom’* components included: curriculum lessons on healthy eating (healthy eating only), healthy eating homework (weight only), role modelling of healthy eating by school community members (healthy eating only), fruit and vegetable breaks (healthy eating only), food growing experiences/school gardens (healthy eating only), healthy cooking programs (healthy eating only), healthy eating embedded into other subjects (healthy eating only), and supporting children to increase their motivation for healthy eating (healthy eating only).

Beneficial *‘healthy food available at school’* components included: choice architecture strategies to encourage students to purchase/select healthier items from canteen/cafeteria (healthy eating only), incentives for selecting/purchasing healthy food and drinks (healthy eating only), school-wide events to promote healthy eating (healthy eating only), providing free fruit and vegetables to children (healthy eating only), free access to cooled or carbonated water (healthy eating only), and providing students free healthy school lunches (healthy eating only).

Beneficial *‘healthy food brought to school’* intervention components included: information to families on healthy eating (weight only).

Beneficial *‘healthy eating outside the school involving families’* components included: interactive healthy eating or cooking programs with students and families (weight only).

##### Physical activity intervention components

A beneficial impact on student physical activity, sedentary behaviour or weight was reported for 8 of the 22 physical activity intervention components.

Beneficial *‘physical activity in the classroom’* intervention components included: curriculum lessons targeting physical activity (weight only) or sedentary behaviour (weight only), physical activity ‘homework’ (weight only), and implementing short activity breaks (weight and MVPA).

Beneficial *‘physical activity outside of the classroom and during break times’* intervention components included: structured break time activities (weight only), and increasing time spent doing physical activity outside of class (weight only).

Beneficial *‘physical activity outside of school or involving* families’ intervention components included physical activity information to parents (weight only), and interactive physical activity programs for students and families outside of school (weight only).

##### Other

A beneficial impact on child weight and healthy eating was also identified for: height and weight screening.

## DISCUSSION

This study sought to develop an inventory of school-based healthy eating, physical activity and obesity prevention intervention componentry targeting children aged 5 to 12 years aligned to global guideline recommendations and consolidate global evidence to explore their effectiveness. This work yielded an inventory of 49 components of interventions targeting healthy eating, physical activity, sedentary behavior, or obesity utilised across 155 primary studies, and spanned each opportunity recommended by guidelines to promote physical activity among school students. Of these we found beneficial impacts on these outcomes for 26 intervention components. The findings demonstrate the breadth of components that have been trialed as part of multicomponent school-based interventions and will facilitate the design of more effective approaches to improve student health.

Beneficial impacts on healthy eating, physical activity, sedentary behaviour, or obesity were found for intervention components across each of the three guideline-aligned opportunities. Components suggested as effective were most common for obesity and healthy eating outcomes. For these components such as “Teacher or student led organised physical activity for students at break time” and “Curriculum learning experiences, curriculum or lessons regarding healthy eating” appear likely to be beneficial. While meta-analyses inclusive of all eligible studies suggests that school based physical activity interventions are, on average, effective in improving MVPA, secondary subgroup analyses examining individual components identified just one to be statistically significant - “short physical activity breaks (e.g. energisers) implemented across the school day”. The findings suggests that comprehensive interventions inclusive of multiple components may be required to improve student activity. Examination of point estimates of secondary subgroup analyses may help to identify a package of components that may maximise potential effects. The absence of existing comprehensive syntheses of school-based studies determining the impact of individual healthy eating and physical activity components limits the ability to compare the findings of this study to previous research. Nonetheless the findings suggest a broad range of intervention components are likely beneficial in addressing obesity related behaviours.

We found broad consistency between the umbrella review and secondary data syntheses approach utilised in this study. For six of the intervention components, both individual systematic reviews were identified, and secondary data analysis was possible. Of the physical activity intervention components, the findings were consistent between the two synthesis methods for three of four intervention components examined. For example, both individual systematic reviews and secondary data analyses consistently found no evidence of effect for active travel and embedding physical activity into non-physical education lessons on child MVPA. Whereas inconsistent evidence was found for short activity breaks, with secondary data analysis suggesting a beneficial impact on time in sedentary behaviour but not MVPA, and systematic reviews reporting no significant impact on either outcome. A similar pattern of results was found for healthy eating intervention components. While differences in the studies included in the syntheses likely explain the inconsistent findings, nonetheless, the findings of systematic reviews designed explicitly to assess the effects of discrete interventions are likely less biassed than secondary subgroup analyses.

Despite the exploratory nature of this study, the findings are particularly useful for policymakers and practitioners given the absence of any comprehensive evidence examining effectiveness of individual components. Identification of those healthy eating and physical activity intervention components that may be the most effective provides important guidance regarding which components should be prioritised within multicomponent interventions to optimise their impact. Importantly the study also highlights those intervention components where there is little evidence of a beneficial impact and could be replaced in multicomponent interventions for other more effective components. The study also highlights a number of healthy eating and physical activity intervention components where their impact on child outcomes have not been synthesised as yet and could be prioritised in future research.

This study has several methodological strengths that enhance its contribution to the existing evidence base. It is the first to systematically consolidate and explore the effectiveness of the discrete intervention components of school-based healthy eating, physical activity and obesity prevention interventions. The intervention component inventory was informed by the most recent and comprehensive systematic reviews targeting the relevant outcomes, to ensure all potential components tested in global studies to date would be captured. Finally, the study employed a rigorous approach to identifying and categorising intervention components, according to internationally recognised guidelines and implementation of iterative coding and consensus process to maximise methodological robustness and validity.

The study also has a number of limitations that should be considered when interpretating the results. Of the primary studies included in the secondary data analysis, 95% were multicomponent interventions and the exploratory subgroup analysis approach adopted to explore effectiveness, cannot isolate the effects of intervention components within multicomponent interventions. As a result, the findings of the secondary data analyses report the effects interventions that included a specific component, and not the effects of that component in isolation. More sophisticated analysis methods that have been developed recently may offer more robust methods for determining the effectiveness of individual intervention components within complex interventions, however these were beyond the scope of the present study. The evidence base used to explore the impact of healthy eating interventions was drawn from an umbrella review and use vote counting, rather than synthesis of primary RCTs. While vote counting can help to capture the frequency of the direction of effect, it doesn’t not quantify the effect size. Additionally, the study considered syntheses of RCTs only, and beneficial impacts of other initiatives synthesised with, or more suited to, non-randomised study designs, or where only single RCTs have been synthesised in reviews conducted to date, were omitted and not considered in determining beneficial impact, and could be investigated in future research.

While the findings of this study provide useful insights , evidence of effectiveness alone is insufficient to make well informed decisions regarding investments in school health initiatives. For example, evidence to decision frameworks suggest policy makers should weigh this with evidence of any unintended adverse effects, considerations of feasibility, cost, and acceptability.^60^ Other factors related to the local context and potential scalability of intervention components should be considered including equity impacts and whether the infrastructure required for implementation is available and the likelihood that any implementation could be sustained. Future research to assess these factors and its integration with the review findings will help better inform the design of impactful and appropriate school-based initiatives.

## Data Availability

All data produced in the present work are contained in the manuscript

## Appendices

**Appendix A:** Search strategy

|  |  |
| --- | --- |
| 1 | (child* or paed* or primary?school* or school?age* or elementary?school* or primary?student or elementary?student or kindergarten*).tw. |
| 2 | (Eating?behavio* or eat* or diet* or fruit* or vegetable* or sugar?sweetened or drink* or beverage* or soft?drink* or water or food* or nutrition or energy?dens* or sugar or energy?rich or portion?size or healthy eating or fast?food* or nutrient?assessment).tw. |
| 3 | (physical* activ* or exercise or physical?fitness or activ* or motion or Movement or Skill or Sedentary?lifestyle or sedentary?time or Sedentary?behavio* or screen?time or sitting?time or inactiv* or sport*).tw. |
| 4 | (body?weight or weight or overweight or obes* or BMI or Body?Mass?Index or waist?circumference or anthropomet* or body?composition or skinfold* or high?weight or fat*).tw. |
| 5 | health behavior/ or body weight/ or overweight/ or obesity/ or body composition/ or waist circumference/ or anthropometry/ or beverages/ or carbonated beverages/ or fruit/ or vegetables/ or skinfold thickness/ |
| 6 | (health?behavio* or behavio?r* or Cogn* or behavio* or learning or attitud* or stigma).tw. |
| 7 | 2 or 3 or 4 or 5 or 6 |
| 8 | "systematic review".pt. or "Systematic Reviews as Topic"/ (((comprehensive* or systematic*) adj3 (review* or literature)) or (meta-analy* or metaanaly*).tw. or ((systematical or systematically) adj2 (review or reviewed reviews)) or (systematically adj identified) or (systematised adj review) or (umbrella adj (review or reviews))).tw. |
| 9 | 1 and 7 and 8 |
| 10 | limit 9 to (english language and yr="2013 - 2023") |

**Appendix B:**
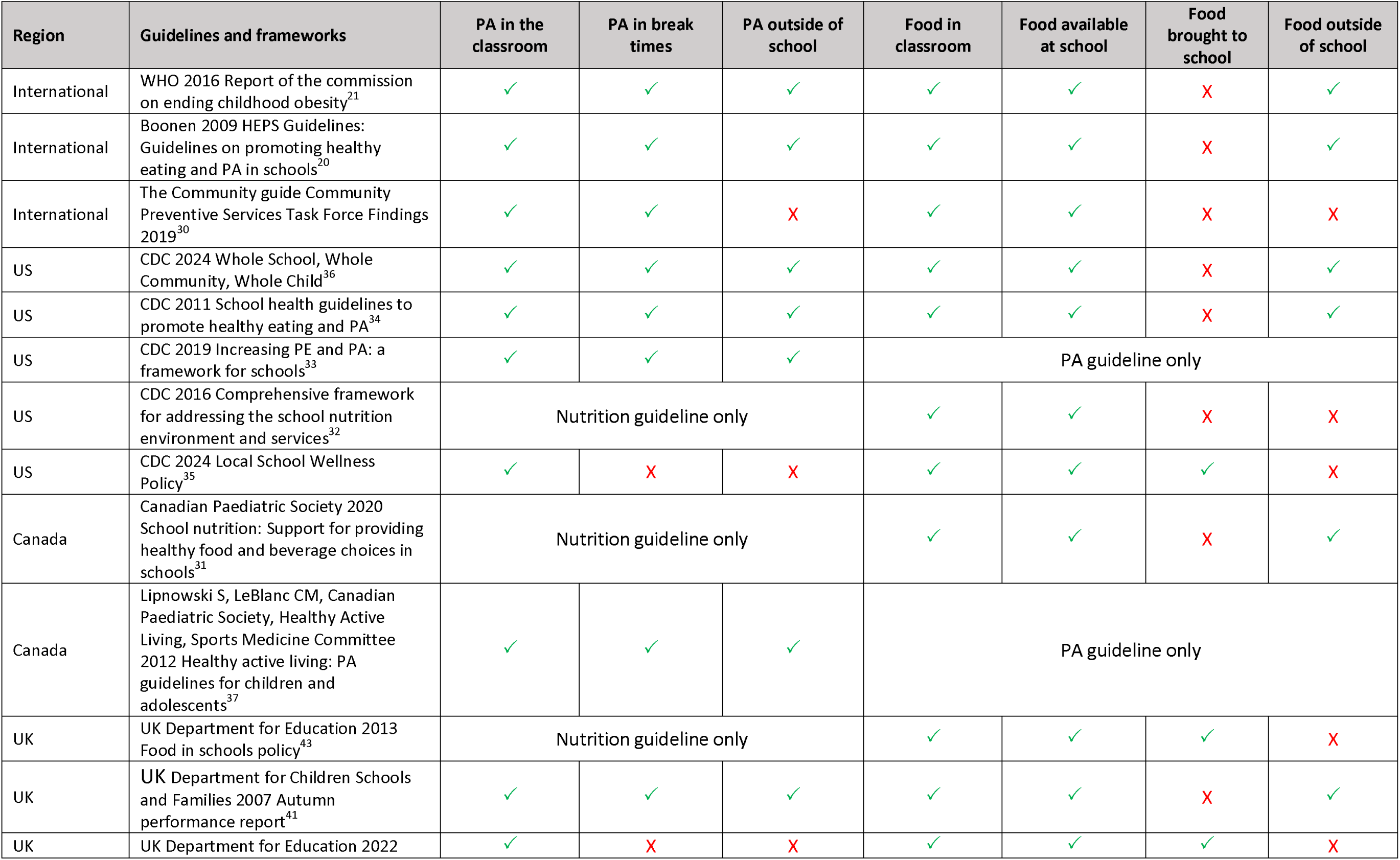
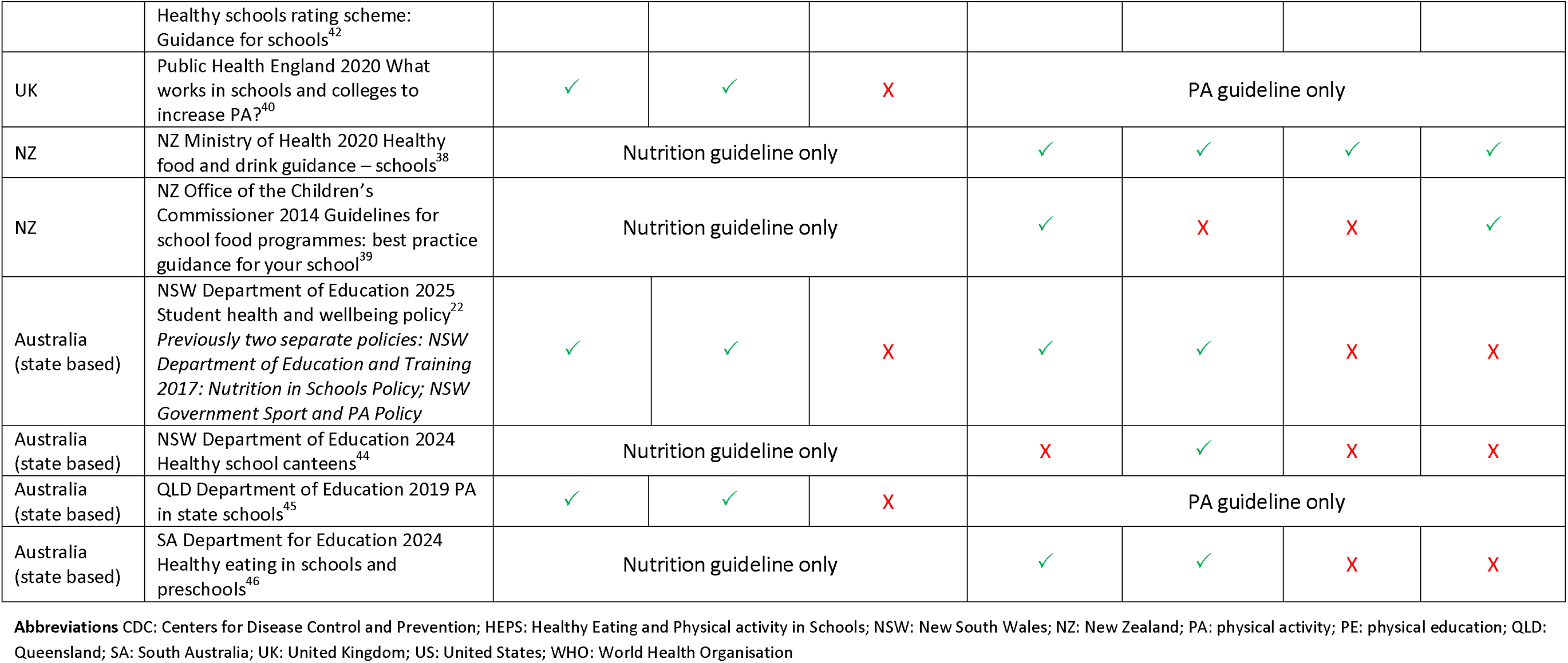
Opportunities for healthy eating and physical activity aligned to global guidelines and frameworks

**Appendix C:** Methodological quality assessment for each systematic review, according to the AMSTAR2 scale

| Reference | 1 | 2 | 3 | 4 | 5 | 6 | 7 | 8 | 9a | 9b | 10 | 11a | 11b | 12 | 13 | 14 | 15 | 16 |
| --- | --- | --- | --- | --- | --- | --- | --- | --- | --- | --- | --- | --- | --- | --- | --- | --- | --- | --- |
| Hodder 2022 | Y | Y | Y | PY | Y | Y | Y | Y | Y | NA | N | Y | NA | Y | Y | Y | Y | Y |
| Neil-Sztramko 2021 | Y | Y | Y | Y | Y | Y | Y | Y | Y | NA | Y | Y | NA | N | Y | Y | Y | Y |
| Lonsdale 2013 | Y | N | N | N | Y | Y | Y | Y | N | NA | N | Y | NA | N | N | Y | Y | Y |
| Masini 2020 | Y | Y | N | N | Y | N | Y | N | Y | NA | N | Y | NA | N | N | Y | N | N |
| Norris 2022 | Y | PY | N | N | Y | Y | Y | PY | Y | NA | N | Y | NA | N | N | Y | Y | Y |
| Nury 2022 | Y | PY | N | PY | Y | Y | Y | Y | Y | NA | Y | Y | NA | Y | Y | Y | Y | Y |
| Ohly 2016 | Y | PY | N | N | Y | Y | Y | PY | N | NA | N | NMA | NA | NMA | Y | N | NMA | Y |
| Peiris 2022 | Y | N | N | N | Y | Y | Y | PY | Y | NA | N | Y | NA | N | Y | Y | Y | Y |
| Pulido-Sanchez 2021 | N | N | N | N | Y | N | Y | N | N | NA | N | NMA | NA | NMA | N | N | NMA | Y |
| Vetter 2020 | Y | N | N | PY | N | Y | Y | N | N | NA | N | NMA | NA | NMA | N | N | NMA | Y |

**Abbreviations:** Y yes; N no; PY partial yes; NMA no meta-analysis conducted

